# Correlation of population mortality of COVID-19 and testing coverage: a comparison among 36 OECD countries and Taiwan

**DOI:** 10.1101/2020.05.27.20113969

**Authors:** Chen Wei, Chien-Chang Lee, Tzu-Chun Hsu, Wan-Ting Hsu, Chang-Chuan Chan, Shyr-Chyr Chen, Chien-Jen Chen

**Affiliations:** Harvard Medical School, Boston, USA; Department of Emergency Medicine, National Taiwan University Hospital, Taipei, Taiwan; Center of Intelligent Healthcare, National Taiwan University Hospital, Taipei, Taiwan; Department of Epidemiology, Harvard TH Chan School of Public Health, Boston, USA; College of Public Health, National Taiwan University, Taipei, Taiwan; Genomics Research Center, Academia Sinica, Taipei, Taiwan; Health Data Science Research Group, National Taiwan University Hospital, Taipei, Taiwan

## Abstract

Although testing is widely regarded as critical to fighting the Covid-19 pandemic, what measure and level of testing best reflects successful infection control remains unresolved. Our aim was to compare the sensitivity of two testing metrics-population testing number and testing coverage-to population mortality outcomes and identify a benchmark for testing adequacy with respect to population mortality and capture of potential disease burden. This ecological study aggregated publicly available data through April 12 on testing and outcomes related to COVID-19 across 36 OECD (Organization for Economic Development) countries and Taiwan. All OECD countries and Taiwan were included in this population-based study as a proxy for countries with highly developed economic and healthcare infrastructure. Spearman correlation coefficients were calculated between the aforementioned metrics and following outcome measures: deaths per 1 million people, case fatality rate, and case proportion of critical illness. Fractional polynomials were used to generate scatter plots to model the relationship between the testing metrics and outcomes. Testing coverage, but not population testing number, was highly correlated with population mortality (r_s_= −0.79, P=5.975e-09 vs r_s_ = − 0.3, P=0.05) and case fatality rate (r_s_= −0.67, P=9.067e-06 vs r_s_= −0.21, P=0.20). A testing coverage threshold of 15-45 signified adequate testing: below 15, testing coverage was associated with exponentially increasing population mortality, whereas above 45, increased testing did not yield significant incremental mortality benefit. Testing coverage was better than population testing number in explaining country performance and can be used as an early and sensitive indicator of testing adequacy and disease burden. This may be particularly useful as countries consider re-opening their economies.

## INTRODUCTIONS

Since the first case of coronavirus disease 2019 (COVID-19) was diagnosed in late December 2019, more than 2.5 million cases and 200,000 deaths have been confirmed worldwide.^1,2^ Without approved drugs or vaccines, aggressive testing, coupled with early isolation of exposed and infected patients, has been most effective at containing the pandemic. Except for countries with small populations however--Iceland being a notable example-mass screening can be difficult.^5^

Absent the capability to test entire populations, determining the level of testing adequate to curb transmission is critical to guiding public health interventions. Both population testing number (tests per million people) and testing coverage (tests per confirmed case) have been cited as appropriate in this regard. This study compared the association between these two metrics and various country-level COVID-19 mortality outcomes, with the goal of identifying the more sensitive predictor of population mortality and deriving a widely applicable benchmark for adequate testing.

## METHODS

Open data through April 12 were collected on COVID-19 testing and outcomes for the 36 OECD (Organization for Economic Cooperation and Development) countries and Taiwan. The Spearman rank correlation test was conducted to evaluate the monotonic relationship between population testing number or testing coverage and several outcome measures, including population mortality rate, case fatality rate, and proportion of critical illness. Scatter plots were generated and fitted by fractional polynomials to model the curvilinear relationship between each testing metric and outcome. A free-knot spline model was used to determine the optimal turning point. The analysis was conducted in R (R Foundation for Statistical Computing, Vienna, Austria) and scatter plots were produced using the package MFP. Comparisons were considered statistically significant for a 2-sided alpha <.05. This study was considered IRB-exempt as it involved analysis of de-identified, publicly available datasets.

## RESULTS

We found that population mortality and case fatality rates were highly correlated with testing coverage (Spearman correlation coefficient (r_s_)=−0.79 and −0.67;P=5.975e-09 and 9.067e-06, respectively) (Figure 1A and 1B). In contrast, the correlation between population testing number and population mortality (r=−0.3;P=0.05) and case fatality rate (r_s_=-0.21;P=0.20) were both weak (Figure 2A and 2B). The proportion of critical cases was moderately correlated with testing coverage and population testing number (r_s_=−0.51 and −0.50;P=0.0017 and 0.0019, respectively) (eFigures 1A and IB).

**Figure 1.**
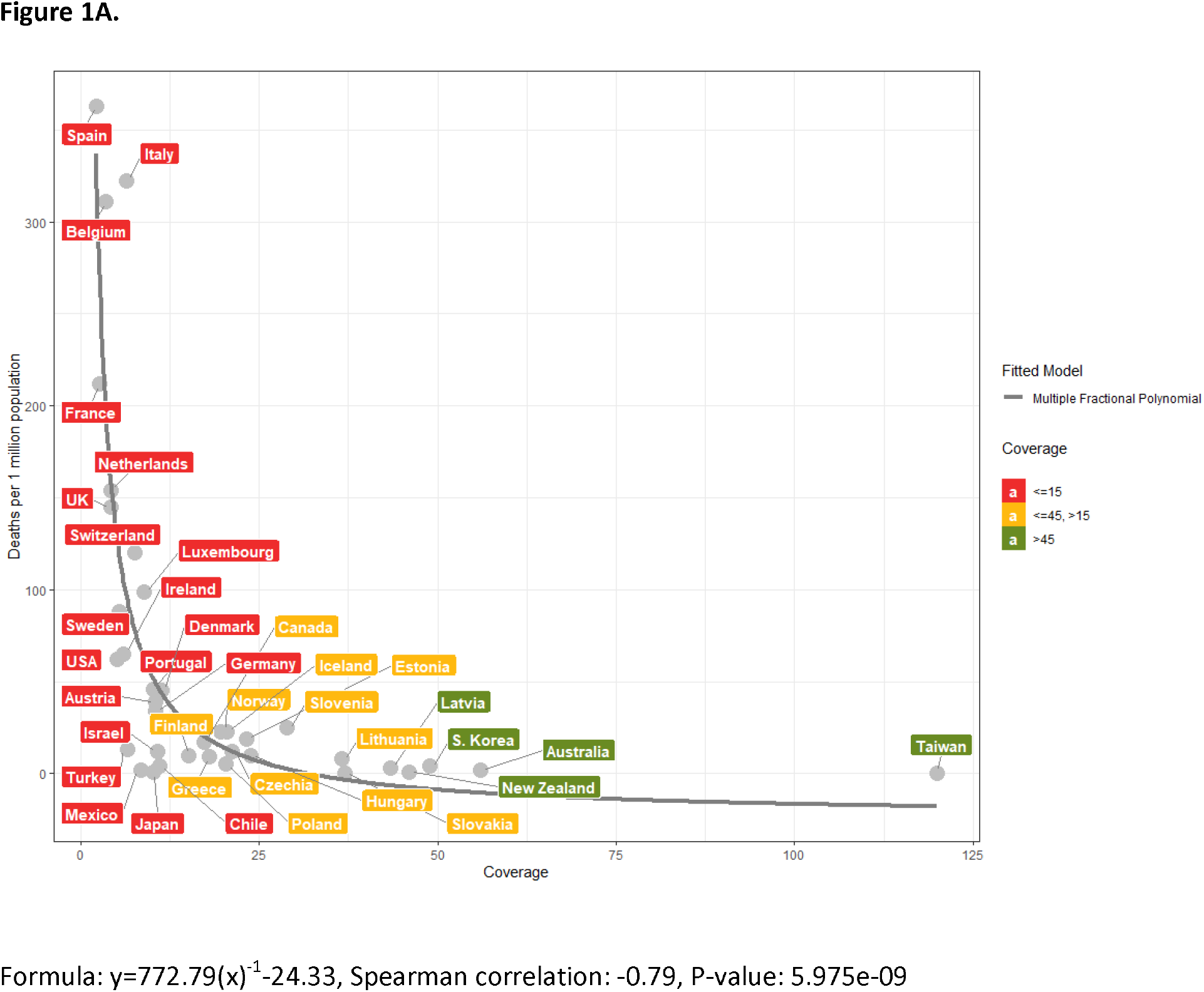
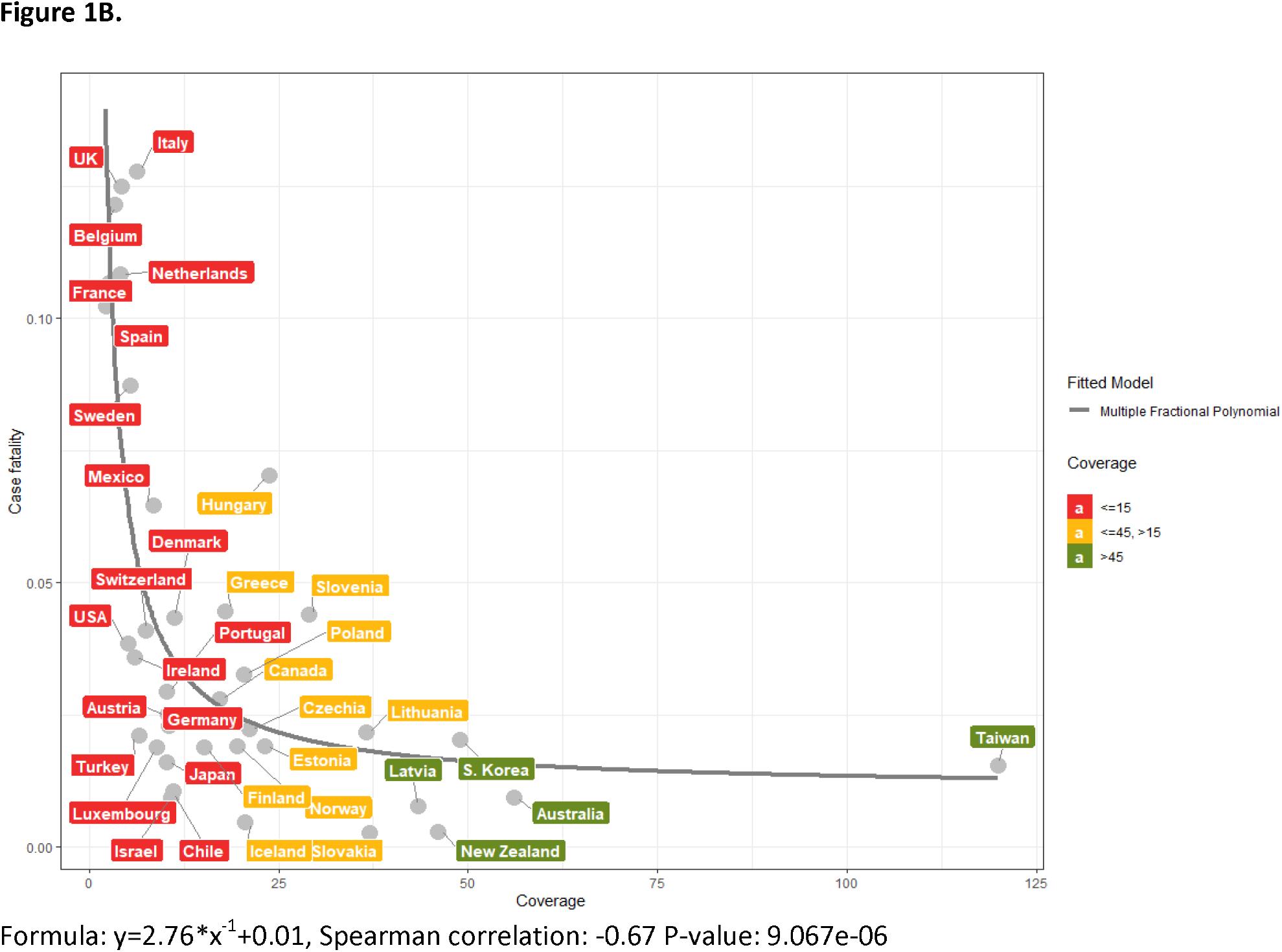
Scatter plots between coverage of tests and outcomes among the 36 OECD countries and Taiwan. The relationship between mortality (per 1 million people) of COVID-19 of 36 OECD countries and Taiwan and coverage of tests **(A)**. The relationship between proportion of case fatalities and coverage of tests **(B)**.

**Figure 2.**
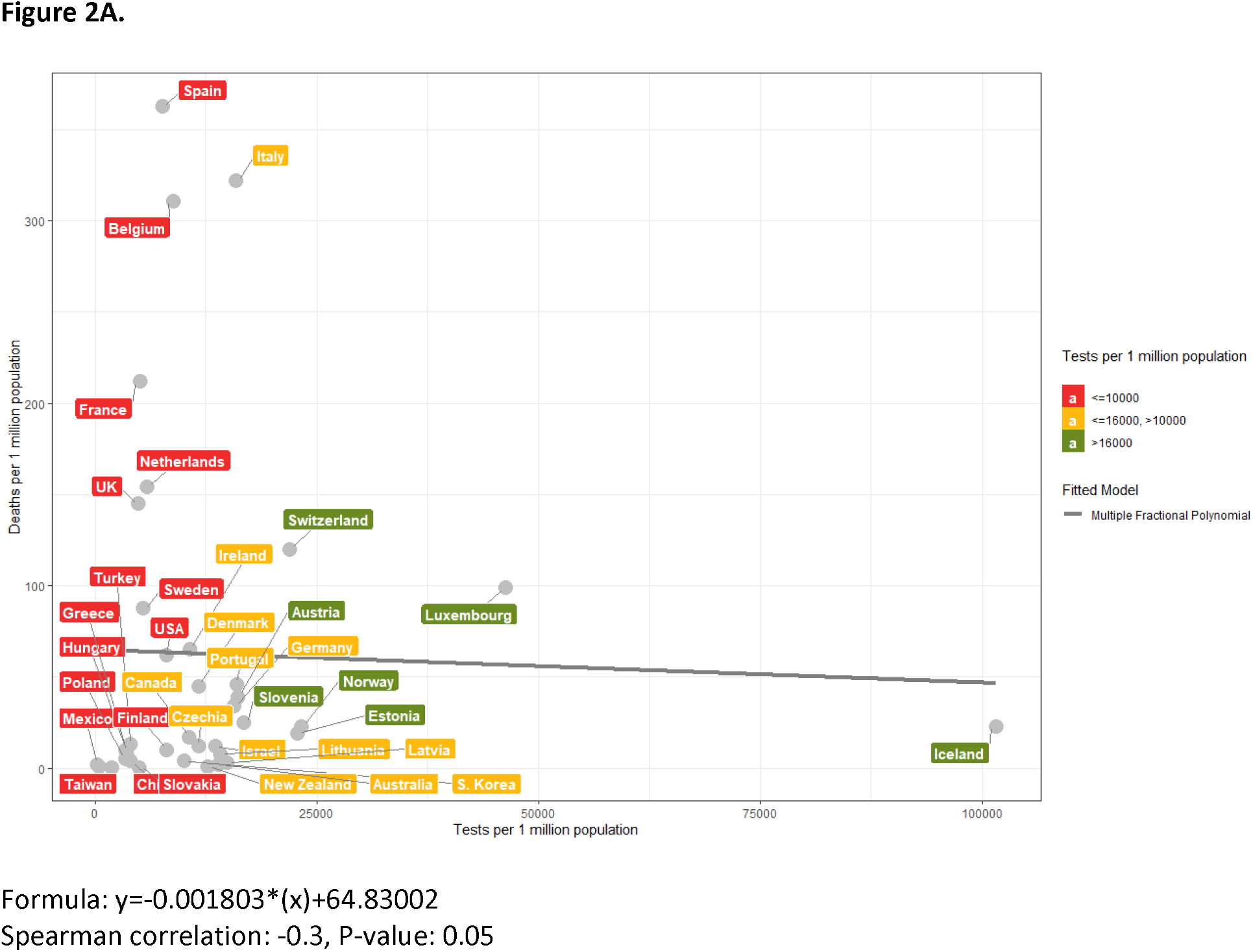
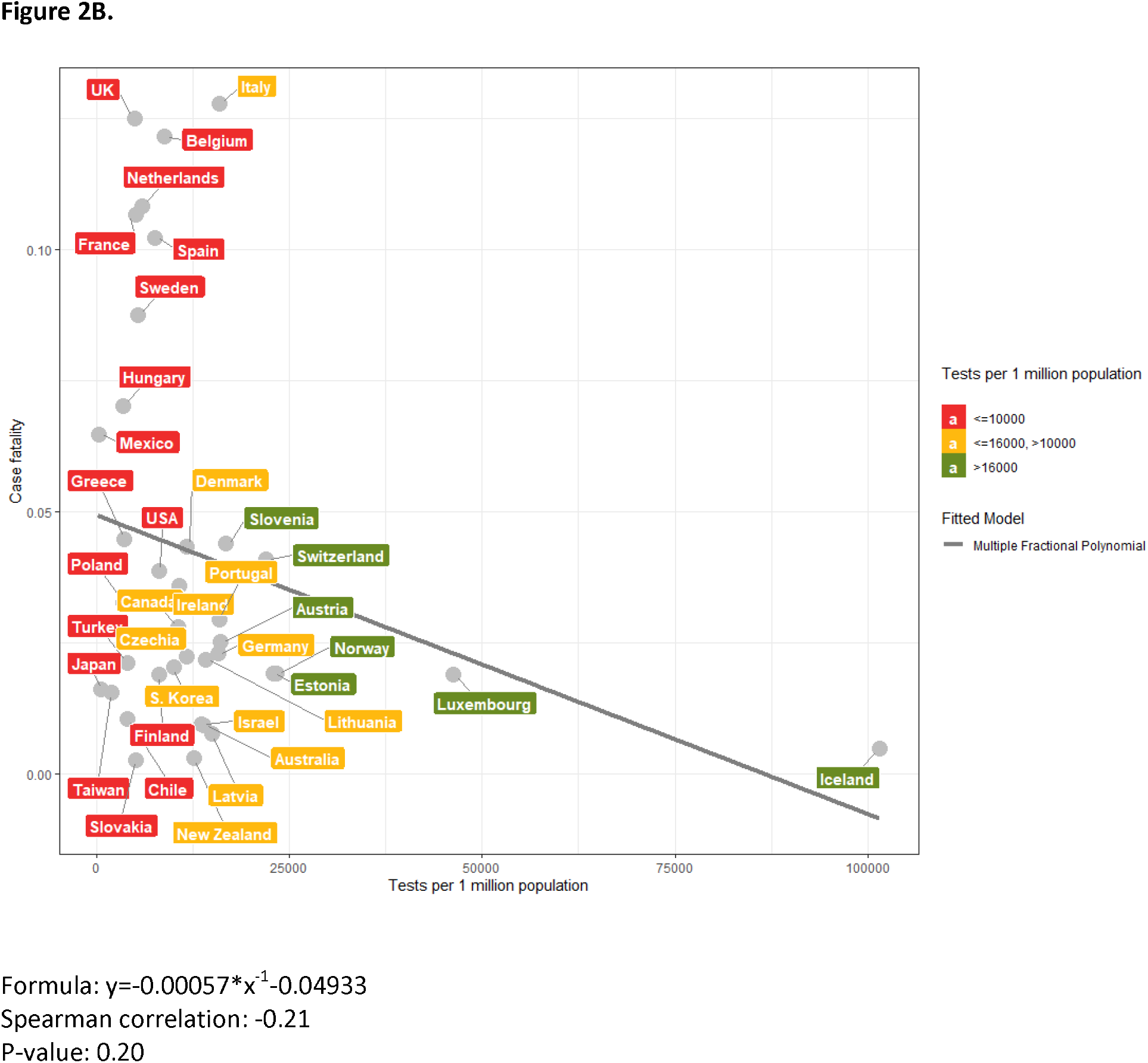
Scatter plots between population testing number and population mortality and case fatality among the 36 OECD countries and Taiwan. The relationship between mortality (per 1 million people) of COVID-19 of 36 OECD countries and Taiwan and number of tests per 1 million people. **(A)**. The relationship between COVID-19 case fatality and number of tests per 1 million people **(B)**.

Detailed testing and outcome measures for each country through April 12 are summarized in Table 1. For the five countries with the lowest testing coverages, Spain (2.1), France (2.6), Belgium (3.4), Netherlands (4.2), and UK (4.2), the population mortality rates were 145,154, 311, 212, and 363 per million people, respectively. In contrast, the five countries with the highest testing coverages, Taiwan (120.0), Australia (56.1), South Korea (49.0), New Zealand (46.0), and Latvia (43.3), reported population mortality rates of 0.3, 2, 4, 0.8, and 3 per million people, respectively.

The inflection point of the mortality curve corresponded to a testing coverage of 15 and flattened after testing coverage exceeded 45 (Figure 1A), and similarly for the case fatality curve (Figure 1B). In contrast, there was only a mildly negative linear relationship between population testing number and population mortality and case fatality, respectively (Figure 2A and 2B).

**Table 1.** COVID-19 disease burden, outcome, and testing number of 36 OECD countries and Taiwan

| Countries | Confirmed cases | Population mortality (deaths/ 1 M) | Population testing number (tests/ 1M) | Critical case ratio | Test positivity rate | Testing coverage |
| --- | --- | --- | --- | --- | --- | --- |
| USA | 533115 | 62 | 8138 | 2.4% | 19.79% | 5.1 |
| Taiwan | 388 | 0.3 | 1954 | NA | 0.83% | 120.0 |
| Australia | 6313 | 2 | 13880 | 2.8% | 1.78% | 56.1 |
| South. Korea | 10512 | 4 | 10038 | 1.9% | 2.04% | 49.0 |
| New Zealand | 1330 | 0.8 | 12684 | 0.6% | 2.17% | 46.0 |
| Latvia | 651 | 3 | 14958 | 0.3% | 2.31% | 43.3 |
| Slovakia | 742 | 0.4 | 5023 | 0.7% | 2.71% | 37.0 |
| Lithuania | 1053 | 8 | 14132 | 1.5% | 2.74% | 36.5 |
| Slovenia | 1205 | 25 | 16764 | 3.5% | 3.46% | 28.9 |
| Hungary | 1410 | 10 | 3471 | 4.9% | 4.20% | 23.8 |
| Estonia | 1309 | 19 | 22878 | 0.9% | 4.31% | 23.2 |
| Czechia | 5905 | 12 | 11684 | 1.6% | 4.72% | 21.2 |
| Iceland | 1689 | 23 | 101497 | 1.3% | 4.88% | 20.5 |
| Poland | 6356 | 5 | 3423 | 8.5% | 4.91% | 20.4 |
| Norway | 6459 | 23 | 23332 | 1.1% | 5.11% | 19.6 |
| Greece | 2081 | 9 | 3583 | 4.4% | 5.57% | 17.9 |
| Canada | 23318 | 17 | 10639 | 3.4% | 5.81% | 17.2 |
| Finland | 2974 | 10 | 8125 | 3.1% | 6.61% | 15.1 |
| Denmark | 5996 | 45 | 11700 | 2.8% | 8.85% | 11.3 |
| Chile | 6927 | 4 | 3995 | 7.7% | 9.07% | 11.0 |
| Israel | 10878 | 12 | 13557 | 1.9% | 9.27% | 10.8 |
| Germany | 125452 | 34 | 15730 | 7.5% | 9.52% | 10.5 |
| Austria | 13945 | 39 | 16086 | 3.7% | 9.63% | 10.4 |
| Japan | 6748 | 0.9 | 544 | 2.0% | 9.81% | 10.2 |
| Portugal | 15987 | 46 | 15966 | 1.5% | 9.82% | 10.2 |
| Luxembourg | 3270 | 99 | 46272 | 1.1% | 11.29% | 8.9 |
| Mexico | 4219 | 2 | 275 | 4.1% | 11.89% | 8.4 |
| Switzerland | 25300 | 120 | 21954 | 3.2% | 13.32% | 7.5 |
| Turkey | 52167 | 13 | 4036 | 3.4% | 15.33% | 6.5 |
| Italy | 152271 | 322 | 15935 | 3.4% | 15.80% | 6.3 |
| Ireland | 8928 | 65 | 10734 | 2.3% | 16.85% | 5.9 |
| Sweden | 10151 | 88 | 5416 | 8.9% | 18.56% | 5.4 |
| UK | 78991 | 145 | 4934 | 2.3% | 23.58% | 4.2 |
| Netherlands | 24413 | 154 | 5926 | 6.4% | 24.04% | 4.2 |
| Belgium | 29647 | 311 | 8814 | 6.2% | 29.02% | 3.4 |
| France | 129654 | 212 | 5114 | 7.7% | 38.84% | 2.6 |
| Spain | 166019 | 363 | 7593 | 8.5% | 46.77% | 2.1 |

## DISCUSSION

Testing has become a paramount concern during the COVID-19 pandemic. To our knowledge, few studies have evaluated how well various testing measures correspond to infection control--in terms of mortality outcomes--and provide benchmarks adequate to curb virus spread. Our investigation demonstrated that testing coverage, but not population testing number, was highly correlated with population mortality and case fatality, and that a threshold of 15-45 indicated testing was adequate to minimize unidentified cases and undetected infection spread.

A critical component of assessing testing adequacy is estimating the magnitude of unmeasured cases in a population. Lachmann and colleagues proposed using a multiplier to convert confirmed case numbers into true case numbers using South Korea’s case fatality rate as a baseline comparator.^6^ Others have also included China using a similar benchmarking approach.^7^ However, these methods can be confounded by country-specific biases in testing and reporting.

Our analysis provides an alternative approach using testing coverage and incorporates data points from 37 countries rather than 1 or 2 countries. Assuming that the SARS-CoV-2 virus is identical across all countries and possesses an intrinsic infection fatality rate, our data suggests that countries with testing coverages of at least 45 need not increase testing further, as higher testing did not correspond with significant decreases in population mortality, likely due to full capture of disease burden. Conversely, countries with testing coverages below 15 may need to ramp up testing and active surveillance.

In addition to informing ideal testing levels, testing coverage can estimate the range of true disease burden in an area by multiplying the confirmed case count by the ratio of 15-45 and the area’s testing coverage. In this way, testing coverage can inform the degree of non-pharmacological intervention (NPI) needed to mitigate community transmission before a potential outbreak worsens. eTable 1 categorizes these interventions into three levels in order of increasing societal and economic costs. As long as the burden of disease in any given area can be reasonably estimated, these measures need not be applied uniformly across an entire country. To this end, testing coverage can be used as a guide to escalate or de-escalate NPIs in dynamic fashion.

Certain conditions, when satisfied, increase the validity of testing coverage for the above indications. Assuming that populations are not infected uniformly, testing accuracy may be subject to stochastic variation as well as systematic sampling bias. As such, its ability to accurately portray disease burden in any area is dependent on access to testing, comprehensiveness of contact tracing, and test sensitivity. Thus far, issues with these criteria have arisen in various degrees across countries, including lagging testing infrastructure, asymptomatic transmission and delayed discovery of cases which complicate contact tracing, and reports of RT-PCR sensitivities ranging from 59-71%.^8,9,10,11^ Despite these limitations however, our analysis showed that testing coverage was still highly correlated with country performance and testing coverage provides additional benefits of low-cost and efficiency.

The results herein should also be interpreted in the context of other limitations. The negative correlation between testing coverage and population mortality does not imply causation, which can only be verified in a prospective interventional study-although there is anecdotal evidence suggesting that early antiviral treatment and/or supportive care may reduce mortality among COVID-19 patients^13^, this may also be due to increased identification of patients with mild disease. In addition, the infection fatality rate of Covid-19 may vary from country-to-country, as has been seen in Italy.^3^ Relevant modifiers include prevalence of risk factors, access to healthcare, robustness of healthcare infrastructure, and population density. Rather than be used in monolithic fashion, testing coverage should therefore be applied in context and with adequate judgment.

In conclusion, we demonstrate the negative curvilinear relationship between testing coverage and COVID-19 population mortality and case fatality rate. Testing coverage can be used as both an indicator of testing adequacy and potential unidentified disease burden, and is most accurate in the context of high healthcare accessibility, comprehensive contact tracing, and testing sensitivity.

## Data Availability

We have provided our data as a supplementary file.

## Publishable disclosure statement of potential conflict of interest

The authors have no conflict of interests to declare.

